# Proteome-wide Mendelian randomization identifies candidate causal proteins for cardiovascular diseases

**DOI:** 10.1101/2023.10.16.23297103

**Authors:** Chen Li, Nicolas De Jay, Supriya Sharma, Katrina A. Catalano, Emily R. Holzinger, Venkatesh Sridharan, Zhaoqing Wang, Lei Zhao, Joseph D. Szustakowski, Ching-Pin Chang, Joseph C. Maranville, Erika M. Kvikstad

**Affiliations:** Informatics and Predictive Sciences, Bristol-Myers Squibb, USA; Immunology and Cardiovascular Thematic Research Center, Bristol-Myers Squibb, USA; Translational Medicine, Bristol-Myers Squibb, USA

## Abstract

**Background:** Cardiovascular diseases (CVD) remain the leading cause of death worldwide, while a lack of clarity on underlying mechanisms has hindered development of novel therapies. Integration of human genetics and proteomics across different ancestries can provide novel, affordable, and systematic approach for target identification and prioritization.

**Methods:** Mendelian randomization approach was applied to unravel causal associations between 2,940 circulating proteins and 21 CVD. Genome-wide summary statistics from the largest genetic mapping of human plasma proteome and meta-analyses on CVD across FinnGen, UK Biobank and Biobank Japan were used. Forward and reverse causation were studied to distinguish respective targets and biomarkers. Genetic instruments for Europeans and East Asians were derived separately and applied to the cardiovascular outcomes in cohorts from corresponding ancestries. We further prioritized drug targets by integrating biological, clinical and population study evidence from cross-database annotations and literature review. Single-cell enrichment analysis and phenome-wide causality scan were performed to further understand target mechanism of action.

**Results:** We found 221 novel candidate causal proteins that impacted risk of one or more CVD through forward MR, and 16 biomarkers whose expression levels were affected by CVD through reverse MR (Bonferroni-adjusted *P*-value <= 0.05). Forward and reverse MR found largely non-overlapping proteins among CVD (only 2 overlapped: LGALS4 and MMP12), suggesting distinct proteomic causes and consequences of CVD. Many of the candidate causal proteins (73.4%) identified are supported by strong literature evidence for a role in immune response and atherosclerotic lesion formation, angiogenesis and vascular remodeling, myogenesis and cardiac progenitor cell differentiation, and energy metabolism. Single cell integration further prioritized ADAM23 for cardiomegaly, PAM for stable angina pectoris and ventricular arrythmia and LPL for peripheral artery disease, whose transcript expression were enriched in cardiomyocytes. Three protein functional groups were highlighted in the phenome-wide scan for their specific enrichment for CVD, including blood coagulation and fibrinolysis, angiogenesis and vascular remodeling, and cell proliferation and myogenesis.

**Conclusions:** Our study identified potential therapeutic targets for CVD and distinguished them from biomarkers due to reverse causation. This study provides human genetics-based evidence of novel candidate genes, a foundational step towards full-scale causal human biology-based drug discovery for CVD.

## Introduction

Recent development in novel therapeutic treatments for cardiovascular disease (CVD) has helped to improve survival and reduce hospitalization of patients.^1,2^ Despite this progress, CVD remains the leading cause of death across ancestries and geographies.^3^ Innovation and investments in developing new therapies for CVD have successfully driven discovery of many novel drugs, including some first- or best-in-class therapies, such as mavacamten, a targeted inhibitor of cardiac myosin, for treatment of patients with obstructive hypertrophic cardiomyopathy;^4^ inhibitors of SGLT2 (dapagliflozin, empagliflozin) to treat heart failure regardless of lek ventricle ejection fraction and diabetes comorbidity,^5–7^ vericiguat, a stimulator of sGC, to treat heart failure patients with reduced ejection fraction;^8^ inhibitors of PCSK9 (alirocumab, evolocumab) for hypercholesterolemia and atherosclerotic CVD.^9,10^ However, in recent years, the number of drugs entering all phases of clinical trials and drugs approved by regulatory bodies (including the US Food and Drug Administration) for CVD has declined, especially when comparing to drugs in other therapeutic areas, such as oncology.^11^ This is possibly due to a relatively higher cost of trials, as cardiovascular trials are often larger in size and longer in duration to manifest primary endpoints that satisfy regulatory requirements.^12^ The challenges confronted in cardiovascular drug development call for new approaches to increase accuracy and efficiency of trials at lower costs, and studies have shown drug targets with robust human genetic evidence are more likely to be successful in clinical trials.^13^

Recent advances in omics technologies provide opportunities to discover novel therapeutic strategies in an unbiased, rapid, and cost-effective manner. High throughput immunoaffinity-based proteomics plamorm is emergingly applied to systematically quantify protein abundance in large cohorts^.14^ Deep profiling of circulating proteins with integration of genomics in a large population cohort allows us to understand proteins associated with disease at an unprecedented scale with the possibility of unraveling novel pathways involved in CVD pathogenesis. Observational studies have shown cross-sectional associations between proteins and certain CVD,^15^ but causal relationships are unclear due to potential confounding effects and reverse causations. Genome-wide association studies (GWAS) of circulating proteins identify genetic variants that regulate protein levels in plasma, which can be used as instrumental variables (IVs) to infer causality under the framework of Mendelian randomization (MR).

Studies have demonstrated the feasibility of MR methodology in identifying protein targets for CVD but are often restricted to a small number of targeted proteins, one or a few CVD and one ancestry population^.16–18^ In this study, we leveraged the largest genetic mapping of circulating proteome that has recently been published as a public data resource, which measures 2,940 proteins in up to 50,000 individuals in UK Biobank^,19^ to systematically identify disease associations across 21 CVD ranging from rare conditions of myocarditis and cardiomyopathy to common conditions of heart failure and ischemic stroke. To distinguish targets of protein expression levels affecting disease risks and biomarkers that vary protein expressions as consequences of diseases, we conducted forward and reverse MR to delineate bi-directional causality. We prioritized newly identified targets and biomarkers with further evidence from cross-database evaluation of pathophysiological function and clinical implication and analyzed potential therapeutic and adverse effects on a wide spectrum of diseases to respectively show opportunities for drug repurposing and caveats for safety issues.

## Methods

### Study Overview

Figure 1 summarizes the overview of our study design – data processing and analysis workflow. The study consists of two parts, the primary analysis, a bi-directional MR that disentangle causal relationships between proteins and CVD, and secondary analyses to strengthen evidence for target prioritization, including single cell integration to assess cell-type enrichment of targets prioritized and differential gene expression between cardiomyopathy and healthy cardiac cell types; manual curation of biological functions and cross-database annotation of protein druggability and disease association; a phenome-wide MR scan to assess pleiotropy and specificity of targets and biomarkers across disease categories.

**Figure 1.**
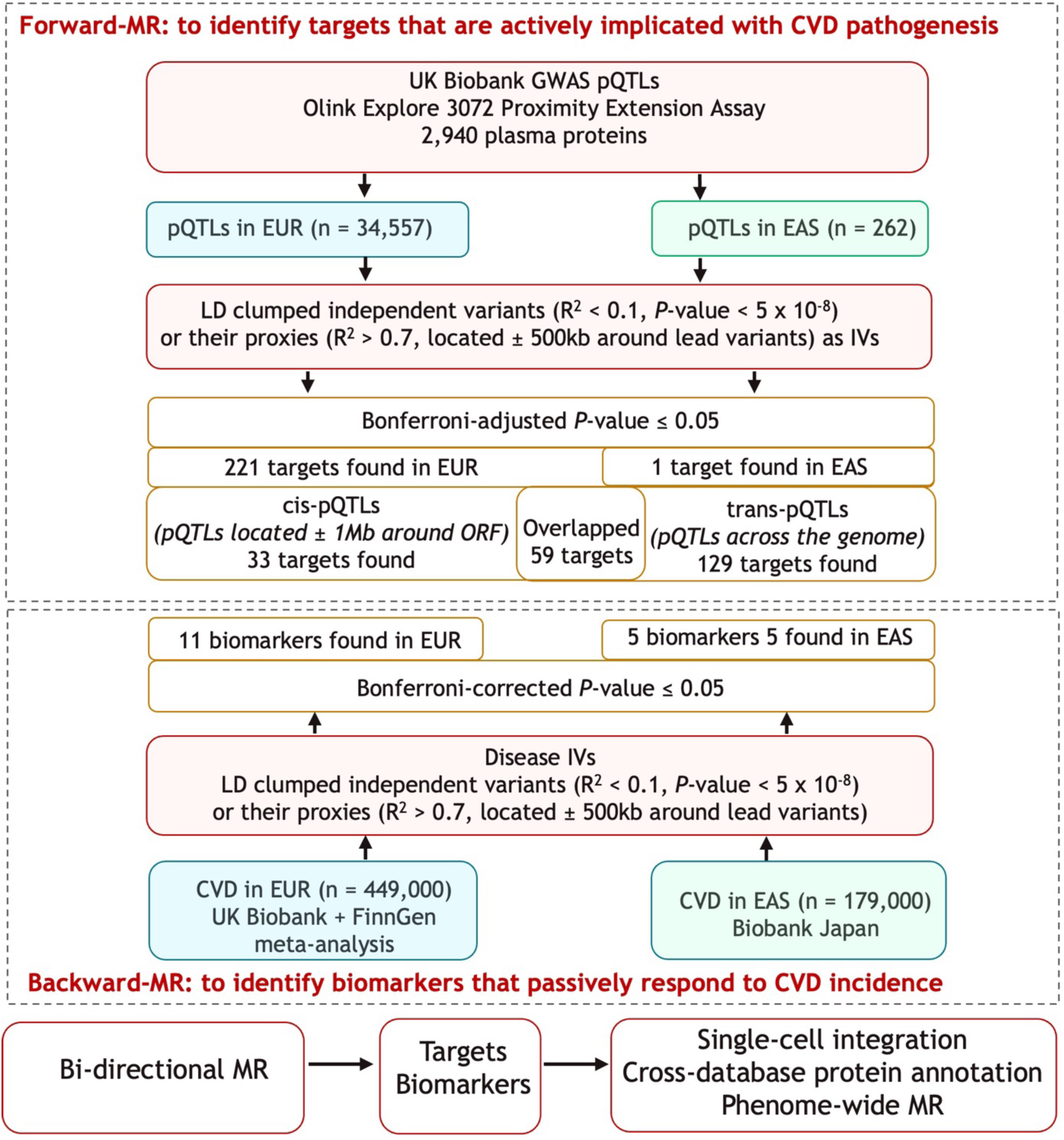
Study Overview. GWAS: genome-wide association study, pQTL: protein quantitative trait loci, EUR: European, EAS: East Asian, LD: linkage disequilibrium, ORF: open reading frame, CVD: cardiovascular disease, IVs: instrumental variables, MR: Mendelian randomization.

### MR analyses

Various methods have been developed to conduct MR with different capabilities in type I and type II error control, and degrees of tolerance to invalid instruments.^20^ We used the inverse variance weighted MR as the primary method because of its conceptually simple design and relatively greater power to detect less significant but true positive results, and other robust methods as sensitivity analyses, including MR Egger, median weighted and mode weighted MR.

#### Forward-MR

Forward MR investigated causal roles of proteins on CVD risk, where proteins are exposures and CVD are outcomes. IVs for proteins are derived from summary statistics from the largest GWAS of plasma proteins, where genetic associations were assessed with 2,940 plasma proteins in ∼54,000 UK Biobank participants, of which 34,557 participants are of European ancestry and 262 participants are East Asians.^19^ Independent lead variants (also known as protein quantitative trait loci, or pQTLs) with *p*-values less than 5 X 10^-8^ were selected as IVs for European and East Asian (Linkage disequilibrium (LD) clumped variants in LD R^2^ < 0.1 with each other and located within 500kb around lead variants) cohorts separately. LD was calculated based on a randomly selected subset of Caucasian (n = 10,000) or all East Asian (n = 2,783) participants in the UK Biobank for Europeans and East Asians, respectively^.21^ For IVs absent from the CVD GWAS datasets, we found the best proxies available (*p*-values < 5 X 10^-8^, in LD R^2^ > 0.7 with index variants, and within 500kb distance from the index variants). We assessed *cis-* and *trans-*pQTLs as IVs separately. *cis-*pQTLs are defined as genetic variants within 1Mb on either side of the start and the stop codon of a protein-coding gene. Proteins that significantly affected CVD risk with Bonferroni-adjusted *P* value < 0.05 (Bonferroni adjust for the total number of proteins and diseases tested) were identified as target candidates and followed-up with downstream analyses of multi-trait colocalization, cross-database protein annotation and phenome-wide MR scan.

#### Reverse-MR

Reverse MR studies protein level changes caused by CVD incidence, where diseases are exposures and proteins are outcomes. IVs for CVD are independent lead variants (LD clumped variants with *p*-value < 5 X 10^-8^, in LD R^2^ < 0.1 between each other and located within 500kb around lead variants) derived from summary statistics of GWAS on 21 CVD where CVD are identified by ICD-10 and phecode in Europeans (UK Biobank and FinnGen meta-analysis, n = 449,000) and East Asians (Biobank Japan, n = 179,000). IV definitions for all CVD are based on the GWAS meta-analyses in respective cohorts^,22^ except for hypertrophic and dilated cardiomyopathy, a more cardiomyopathy-focused GWAS with greater statistical power was used for Europeans for these 2 relatively rare cardiovascular disorders^.23^ Ancestry-specific LD was estimated in corresponding cohorts as illustrated in the Forward-MR analysis. For IVs absent from the proteomics GWAS datasets, we found the best proxies available using the same criteria as described in the forward-MR analysis. Proteins that are significantly affected by CVD (Bonferroni-adjusted *P* value < 0.05, Bonferroni adjust for the total number of proteins and diseases tested) are identified as potential biomarkers and compared against significant results from the forward-MR to rule out reverse causation.

### Cross-database protein annotation

Protein biological functions are annotated with cross-referencing multiple databases, including UniProt and RefSeq. We also annotated protein druggability by interrogating Drug-Gene-Interaction (DGI) database,^24^ ChEMBL v33,^25^ PharmGKB^,26^ and DrugCentral Postgres v14.5^27^ databases to find therapeutic targets of approved or under clinical development. To evaluate protein associations with diseases, we queried GWAS Catalog^28^ for population-level evidence, as well as ClinVar^29^ and ClinGen^30^ for clinically suggested implications.

### Phenome-wide MR scan

We ran MR to disentangle bi-directional causal relationships between 2,940 proteins and 159 disease outcomes in European and East Asian cohorts separately. For East Asian cohort, an additional 38 quantitative biomarkers and 23 medication usage phenotypes were tested. Diseases were defined by ICD-10 and phecode by a previous GWAS meta-analysis in UK Biobank, FinnGen and Biobank Japan, where detailed description of the 3 cohorts and disease definitions are shown^.22^ The diseases cover a wide range of categories, with an average of 8 diseases representing each category. Significant results from forward- and reverse-MR are followed up with the phenome-wide MR scan to detect off-targets effects and opportunities for drug repurposing. Diseases that have ≥ 50 cases were included in the forward- and reverse-MR analyses and at least one genome-wide significant locus in the reverse-MR analysis. Enrichment was assessed via the over representation analysis to determine whether a protein functional group (G) was over-represented in a disease category (D), and enrichment *P* value was calculated under hypergeometric distribution using the formula: 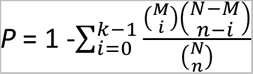, where N is the product of the total number of diseases across disease categories and the total number of proteins in G, M is the product of the total number of diseases in D and the total number of proteins in G, n is the number of significant protein signals in G across disease categories, and k is the number of significant protein signals in G and D. Enrichment significance threshold is Bonferroni-corrected for the total number of protein functional groups and the total number of disease categories.

### Single-cell analysis

To link circulating proteins to proteins enriched in cell types originating in the heart tissues or demonstrate specificity for cardiomyopathy *vs* healthy hearts, we analyzed cell type enrichment for significant genetic signals from MR analyses using single-cell RNA sequencing data from left ventricular samples and assessed differential gene expression between cardiomyopathy and healthy hearts.

#### Single-cell RNA sequencing data cura;on

We curated publicly available single-cell RNA sequencing data from a recently published cohort of 42 human left ventricular samples (11 DCM, 15 HCM, 16 non-failing hearts).^31^ Cell and cell-type metadata, and gene counts adjusted for ambient RNA were retrieved from the Broad Institute’s Single Cell Portal (accession SCP1303). Cell-type enrichment and differential gene expression data were retrieved from figure source data or supplementary tables of the corresponding publication.

#### Cell-type enrichment

To determine whether a gene exhibited significantly enriched expression in one or more cell types, pseudobulk profiles were generated from healthy and patient donors.^31^ A gene was considered to be enriched for a cell type if it satisfies the following criteria: (1) the gene expression was > 4X higher for the cell type compared to the other cell types (one *versus* rest) with FDR_BH_-adjusted *P*-value < 0.01, (2) at least 25% of nuclei in the cell type expressed the gene, and (3) a classifier trained on the gene expression predicted whether a nucleus belongs to the cell type with AUC > 0.6. For visualization, we included genes if their expression were significantly enriched in at least one cell type and sorted them according to (1) the cell type in which nuclei displayed the highest average expression, (2) the average Gini coefficient calculated from the average nuclei expression and the percentage of nuclei expressing that gene.

#### Differential gene expression between cardiomyopathy and healthy hearts

Chaffin et al. analyzed pseudobulk profiles using a limma-voom model regressing gene expression in each cell type on a disease group (HCM vs control and DCM vs control) adjusted for age and sex. A gene was differentially expressed if the expression was 50% higher in the cardiomyopathy than non-failing cardiac cell types with a FDR_BH_-adjusted *P*-value < 0.01.^31^

### Analysis software

All analyses were performed using the statistical software R (Version 4.1.3). We used the PLINK 1.9 beta and 2.0 alpha (https://www.cog-genomics.org/plink/) to generate LD matrix for each ancestry and perform LD clumping, the TwoSampleMR R package (https://github.com/MRCIEU/TwoSampleMR) to perform MR analyses^,32^ and the Seurat (https://satijalab.org/seurat/) and the edgeR R packages to perform single cell analyses.^33,34^

### Ethical Statement

The MR analyses were based on publicly available GWAS summary statistics and ancestry-specific LD matrices were estimated using UK Biobank individual-level data (Applications 26041 and 65851). The included GWAS all received informed consent from study participants and have been approved by pertinent local institutional review board.

## Results

### Target discovery with forward-MR – causal effects of proteins on each CVD risk

In the forward-MR analysis in European ancestry, 221 proteins significantly affect at least one CVD risk (*P*-value < 1.27 X 10^-7^ or 2.09 X 10^-7^ using *trans*- or *cis*-pQTL as instrumental variables, respectively), which spanned multiple Olink panels, including cardiometabolic, inflammation, neurology, and oncology panels (Figure 2). Up to 60% significant proteins affected only one CVD, while several proteins were causally associated with a highly pleiotropic set of diseases across CVD (Figure 2), such as LPA with angina pectoris, atrial fibrillation, ventricular arrhythmia, myocardial infarction, peripheral arterial disease, ischemic stroke, cardiac valvular disease, and cardiomegaly, possibly through dual pathological auributes of lipoprotein(a) in procoagulant effects of apo(a) and atherogenic and proinflammatory effects of oxidized apoB-related phospholipids.^35^ Studies suggested LPA as an emerging therapeutic target for reducing lipoprotein(a) levels, thereby lowering risks of atherosclerotic CVD independent of lowering low-density lipoprotein cholesterol levels,^36^ with positive results reported by several ongoing clinical trials^.37^ Most of the targets identified were novel, while several established targets for cardiovascular complications were also been found, including PCSK9 for myocardial infarction, stroke, and coronary revascularization, AtiGPTL3 for reducing low-density lipoprotein cholesterol, and ECE1 for congestive heart failure and hypertension (Table S1). Many of the targets (62%) identified are supported by strong literature evidence for a role in immune response and atherosclerotic lesion formation, angiogenesis, and vascular remodeling, myogenesis and cardiac progenitor cell differentiation, and energy metabolism (Table S2). Full forward MR results are provided in Table S4.

**Figure 2.**
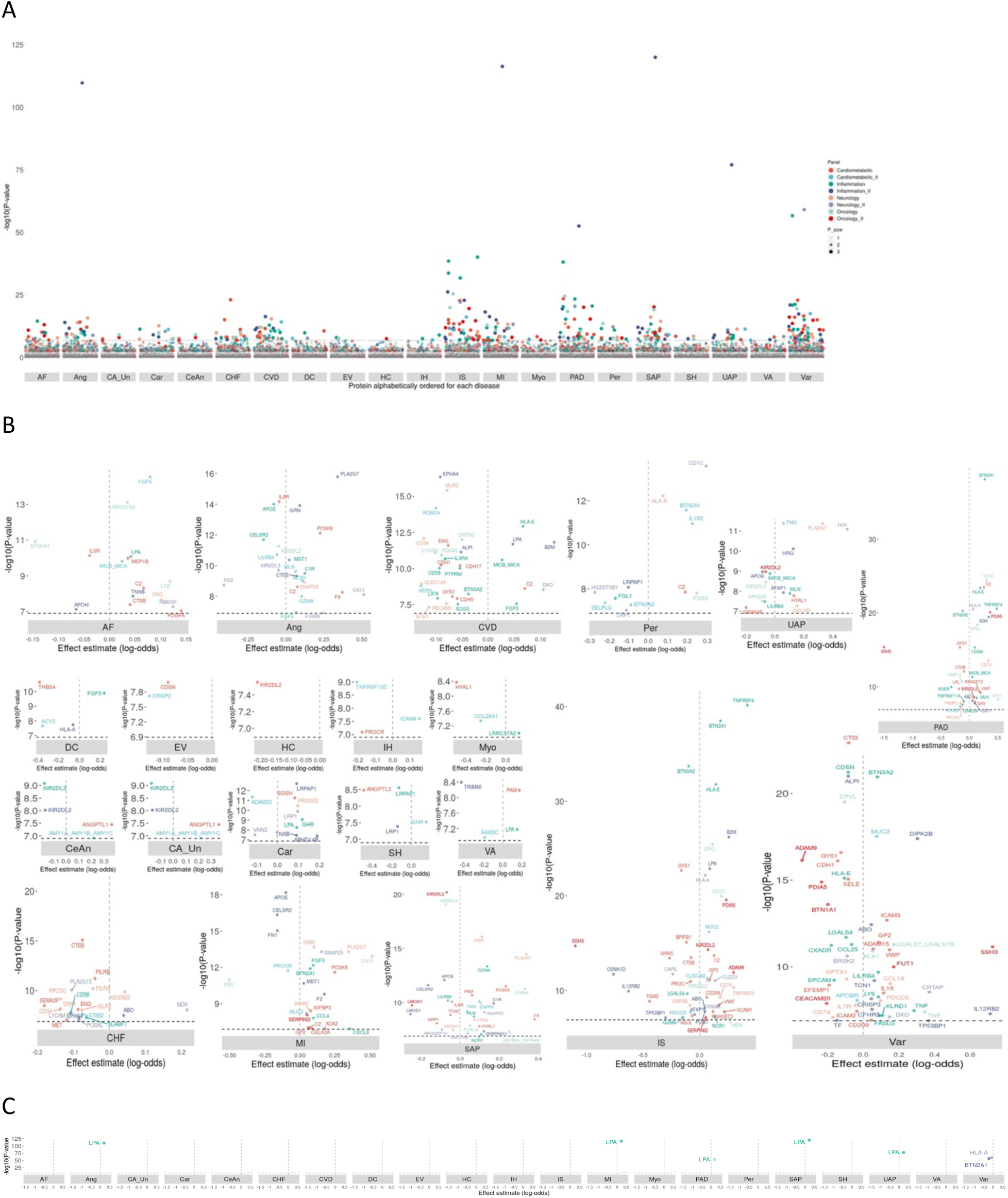
Discovery of targets – circula:ng proteins that are implicated with pathogenesis of cardiovascular diseases (CVD). Forward-MR results using trans-pQTLs as instrumental variables and inverse variance weighted method i n the European ancestry are shown. A. Proteome-wide ManhaQan plot – causal association of each protein with each CVD. -log_10_(*P* -value) was ploQed as Y-axis and protein name in alphabetical order as X-axis for each CVD. Color labels the Olink Panel that each protein belongs to and size labels significant levels, which was categorized into 3 gro ups: groups 1,2 and 3 corresponding to *P*-value ≥ FDR_BH_ threshold of 0.05, *P* -value between Bonferroni-corrected a nd FDR_BH_ thresholds of 0.05 and *P* -value < Bonferroni-corrected threshold of 0.05. Significant causal associations fo r each CVD were shown in the panels B (*P* -value > 1 X 10^-50^) and C (*P* -value ≤ 1 X 10^-50^). Effect coefficient β was plot ted as X-axis and -log_10_(*P* -value) as Y-axis. Dashed vertical line indicates β = 0. Each dot was labeled with protein na me abbreviation colored by the corresponding protein Olink Panel. Ang: Angina pectoris, AF: Atrial fluQer/fibrillation, CVD: Cardiac valvular disease, Car: Cardiomegaly, CeAn: Cerebral aneurysm, CHF: Chronic heart failure, DC: Dilated cardiomyopathy, EV: Esophageal varix, HC: Hypertrophic cardiomyopathy, IH: Intracerebral hemorrhage, IS: Ischemic stroke, MI: Myocardial infarction, Myo: Myocarditis, Per: Pericarditis, PAD: Peripheral arterial disease, SAP: Stable angina pectoris, SH: Subarachnoid hemorrhage, CA_Un: Unruptured cerebral aneurysm, UAP: Unstable angina pectoris, Var: Varicose, VA: Ventricular arrhythmia.

### Biomarker discovery with reverse-MR – causal effects of CVD on each protein level

We identified 16 biomarkers in total, among which 5 were from the East Asian analyses (ERBB3, SIRT5, CXCL13, SUSD5, TTR) shown in Table S5 and Figure 3. Of note, ERBB3, a member of membrane-bound tyrosine kinase receptor family that activates cell proliferation and differentiation, found only in the East Asian population, has recently been shown to have cardioprotective effects on cardiomyocyte survival and angiogenesis under stress^.38^ Genetic polymorphisms of ERBB3 were reported to be associated with coronary artery disease in Han and Uygur ancestries of China,^39^ and circulating protein levels with overweight-related hypertension in the same population.^40^ Only 2 biomarkers (LGALS4, MMP12) were overlapped with the targets derived from the forward-MR analyses (Table S5), suggesting distinct roles of proteins on CVD pathogenesis. We also found NPPB, the gene encoding preprohormone (propro-B-type natriuretic peptide, preproBNP) in cardiomyocytes, as a biomarker specifically to dilated cardiomyopathy. The preproBNP is cleaved into proBNP, and then further processed into 2 circulating fragments -- the biologically active BNP and the inactive N-terminal proBNP, both of which are routinely used in clinical diagnosis and treatment management in heart failure. Studies have suggested that BNP may play different roles in heart failure with preserved and reduced ejection fraction (HFpEF and HFrEF, respectively)^,41^ and BNP or NT-proBNP were less effective biomarkers for HFpEF due to their average lower values in HFpEF than in HFrEF, which dropped into a normal range in some patients^.42^ Our finding is consistent with these studies that BNP-related biomarkers are mainly indicative for heart diseases with reduced left ventricle ejection fraction.

**Figure 3.**
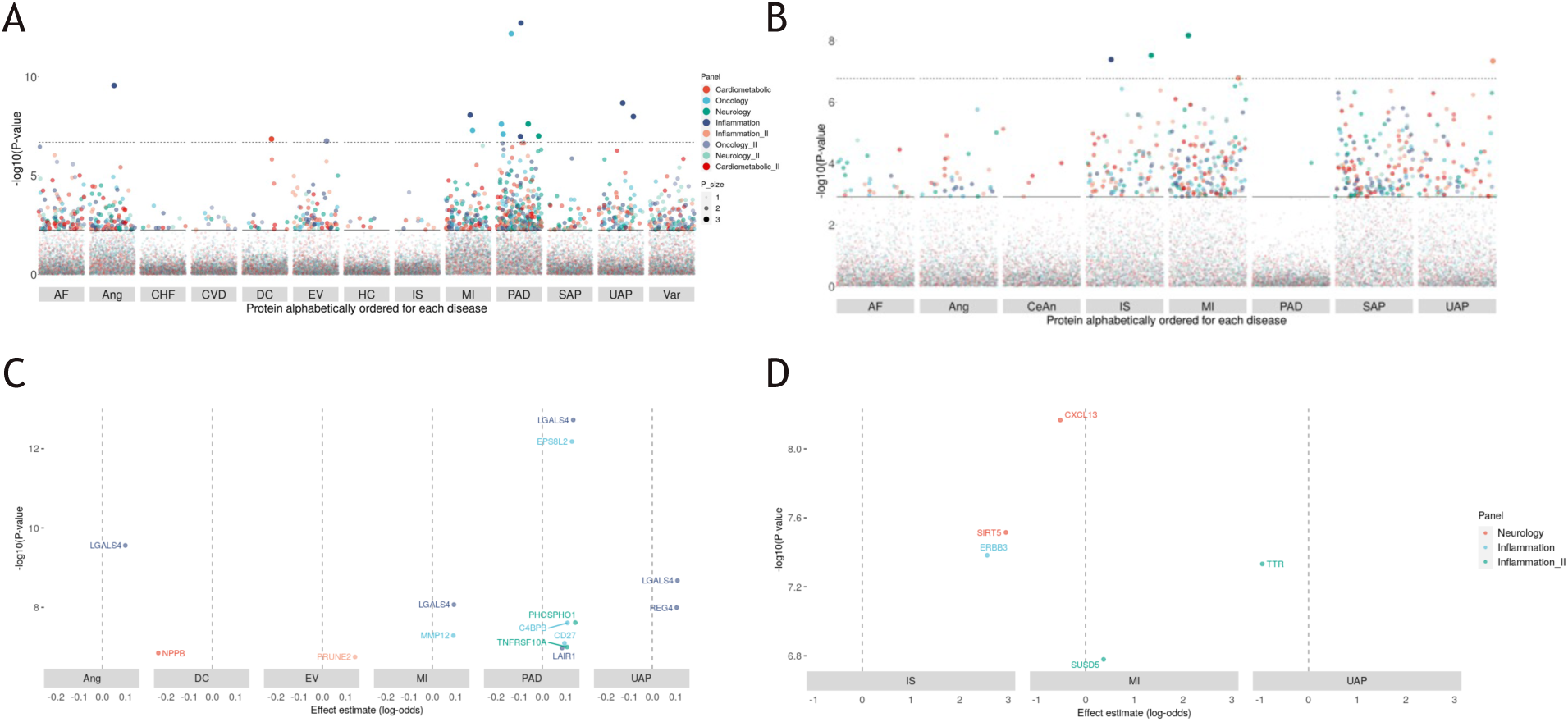
Discovery of biomarkers – circula:ng proteins that respond to cardiovascular disease (CVD) development. Reverse-MR results using inverse variance weighted method across ancestries are shown. Proteome-wide ManhaQan plot – causal effect of each CVD on each protein in the panel A (European) and B (East Asian) ancestries. CVD that had at least one genome-wide significant variant association were shown. Significant causal associations for each CVD were shown in the panels C (European) and D (East Asian). CVD that had causal effects on at least one protein were shown. Figures were represented in the same manner as in the Figure 2. AF: Atrial fluQer/fibrillation, Ang: Angina pectoris, CHF: Chronic heart failure, CVD: Cardiac valvular disease, CeAn: Cerebral aneurysm, DC: Dilated cardiomyopathy, EV: Esophageal varix, HC: Hypertrophic cardiomyopathy, IS: Ischemic stroke, MI: Myocardial infarction, PAD: Peripheral arterial disease, SAP: Stable angina pectoris, UAP: Unstable angina pectoris, Var: Varicose.

### Phenome-wide MR screening for specificity and pleiotropy of targets and biomarkers

We expanded our analysis to evaluate all proteins and all diseases in the forward and reverse MR across the two ancestries to assess pleiotropy and specificity of targets and biomarkers that trigger or respond to one or multiple diseases (Table S6). For targets, 38 out of 190 targets (trans-forward-MR) were specific to one disease and 51 to one disease category; for biomarkers, 4 out of 16 biomarkers were specific to one disease and the diseases were all in the cardiovascular category, such as NPPB specific to dilated cardiomyopathy. Several functional groups showed significant enrichment for their causality on CVD and not the other disease categories, including blood coagulation and fibrinolysis (enrichment P-value = 3.54 X 10^-7^), angiogenesis and vascular remodeling (2.29 X 10^-6^), and cell proliferation and myogenesis (2.38 X 10^-5^). Full enrichment results of protein functional group across disease categories are shown in Table S7. To assess pleiotropy, 37.8% (14 out of 37) proteins in the functional group of immune and inflammatory response exerted significant causal effects on at least 20 different diseases across disease categories, whereas in the other top 10 largest functional groups, there was ≤ 1 protein implicated with ≥ 20 diseases. The pleiotropic signature of the immune and inflammatory response group was consistent when altering the pleiotropy threshold to 15, 10 or 5 proteins (Table S8), suggesting involvement of this functional group in the pathogenesis of a variety of diseases. Apart from this group, several other proteins of different functional groups showed extensive pleiotropy, such as CTSB and MST1 in regulating autophagy and apoptosis, and TNXB in mediating cell-cell and cell-matrix interaction. Biomarkers were associated with a smaller number of diseases on average than targets (mean [SD]: 4.56 [3.29] vs 8.71 [12.28]), and they were not enriched for any disease categories (Figure 4).

**Figure 4.**
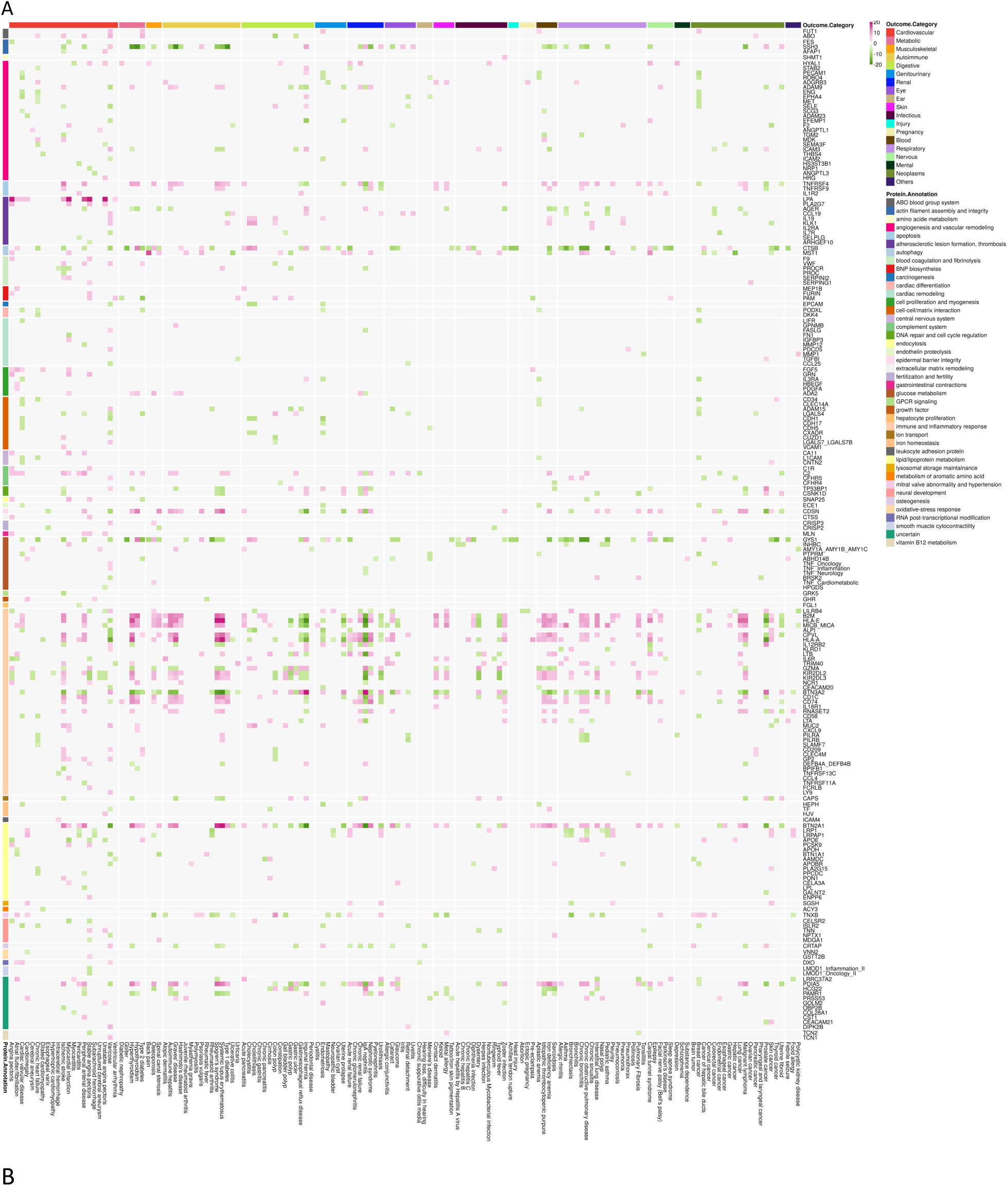
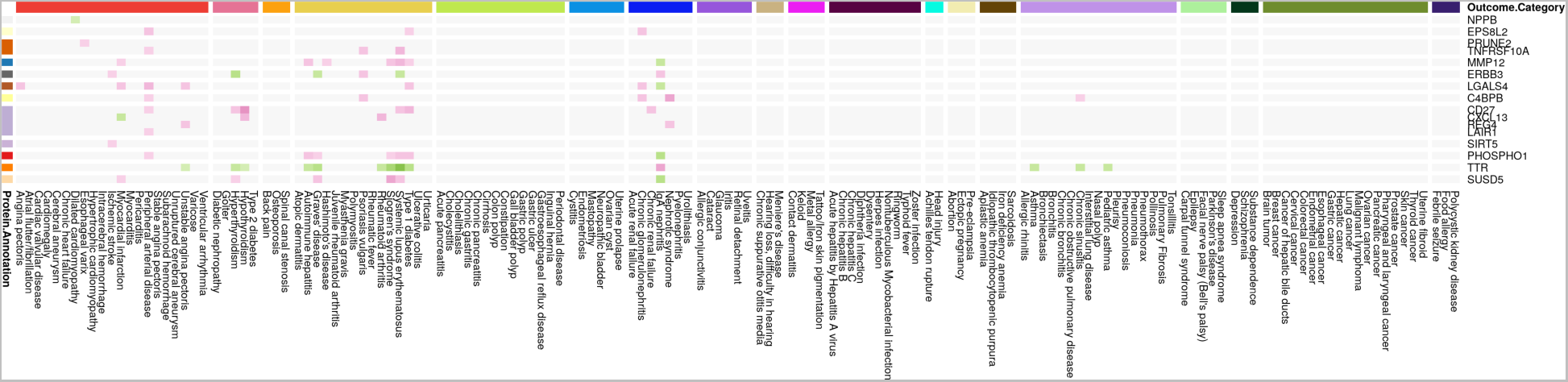
Phenome-wide MR scan to evaluate target and biomarker specificity and pleiotropy. Heatmap representation of Z-scores from A. forward-MR results using trans-pQTLs as instrumental variables and B. reverse-MR results, with inverse variance weighted method in the European ancestry. For Z-scores of absolute values > 20, they are truncated to a maximum absolute value of 20. The color gradient, from green (negative) to red (positive), illustrates direction and magnitude of causal associations between proteins (Y-axis) and diseases (X-axis). Diseases and proteins are grouped by their different categories and labeled with different colors on the top and the lel-side of the heatmap, respectively.

### Single-cell integration elucidates cell context-dependent mechanisms of targets

To gain insights into possible cell context-dependent mechanisms of these targets and biomarkers, we examined their gene expression pauerns along major cardiac cell types of the left ventricle identified in a recently published single-cell late-stage DCM and HCM heart atlas.^31^ Fifty-four out of 235 proteins (22%) had significantly enriched gene expression in at least one major cardiac cell population (Figure 5A). The enriched cell types of each protein were in line with its biological functions (Figure 5B). For example, ADAM23, a protein involved in cell-cell and cell-matrix interactions and cardiac remodeling had significantly elevated expression in cardiomyocytes relative to other cell types. Cardiac-specific conditional knockout of Adam23 in mice exhibited cardiac hypertrophy and fibrosis, whereas transgenic mice overexpressing Adam23 in the heart exhibited reduced cardiac hypertrophy in response to pressure overload.^43^ ADAM23 was a significant causal signal in both cis- and trans-MR, specifically for cardiomegaly, while other paralogs in the ADAM protein family, such as ADAM15 and ADAM9, were more pleiotropic, broadly affecting a highly heterogeneous spectrum of diseases. *PAM*, a known marker of cardiomyocytes encoding the major atrial membrane protein involved in proANP containing secretory granule biosynthesis, was enriched in cardiomyocytes. Similarly, *LPL*, which encodes a key enzyme in hydrolysis of triglyceride and catabolism of triglyceride-rich lipoprotein, was significantly elevated in cardiomyocytes and adipocytes. Mislocalization of LPL to the cell surface of cardiomyocytes has been associated with cardiomyopathy pathogenesis in mice.^44^ Our findings suggest that cell-type-dependent mechanisms of targets emerge as compelling focal points for potential therapeutic hypotheses development.

**Figure 5.**
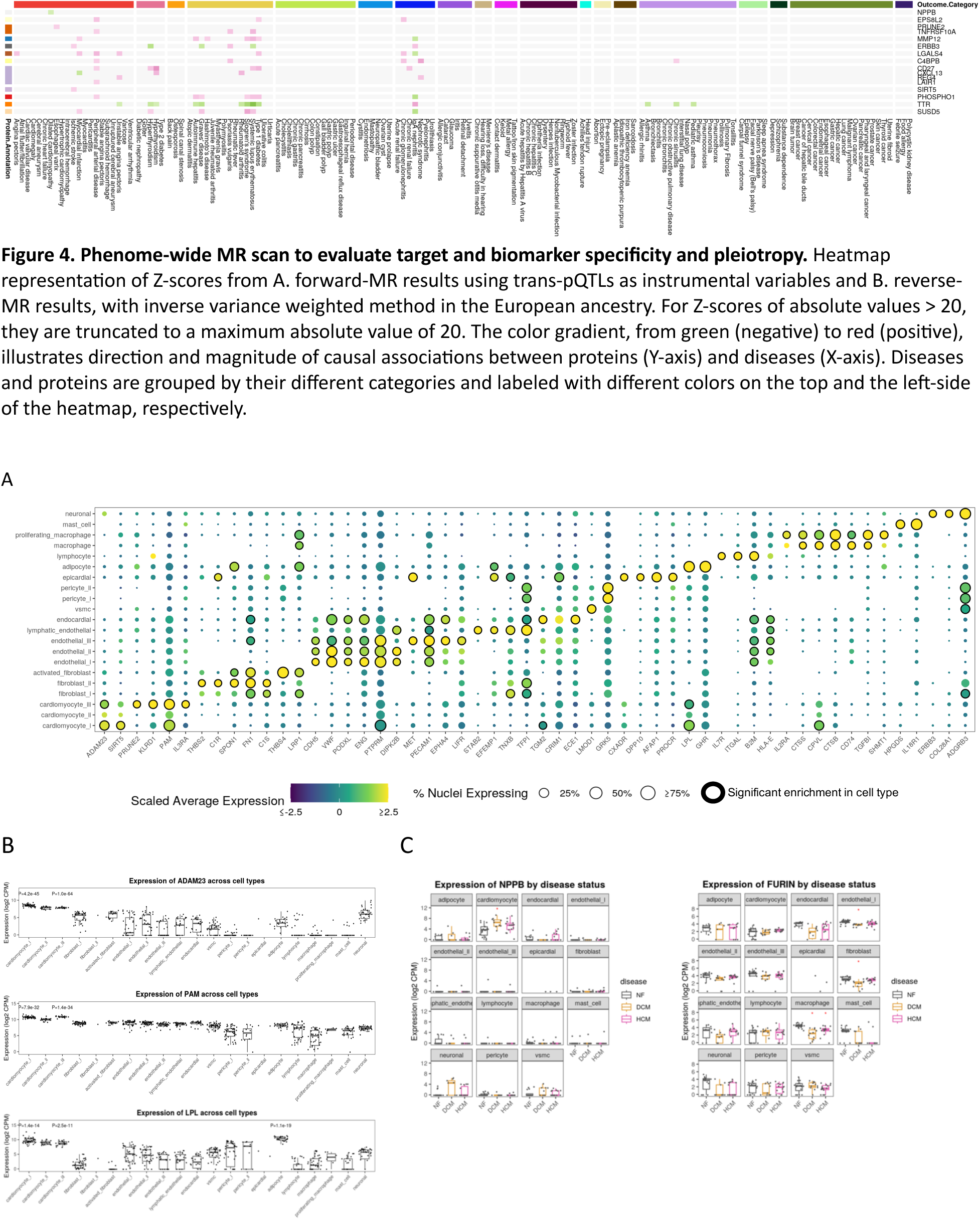
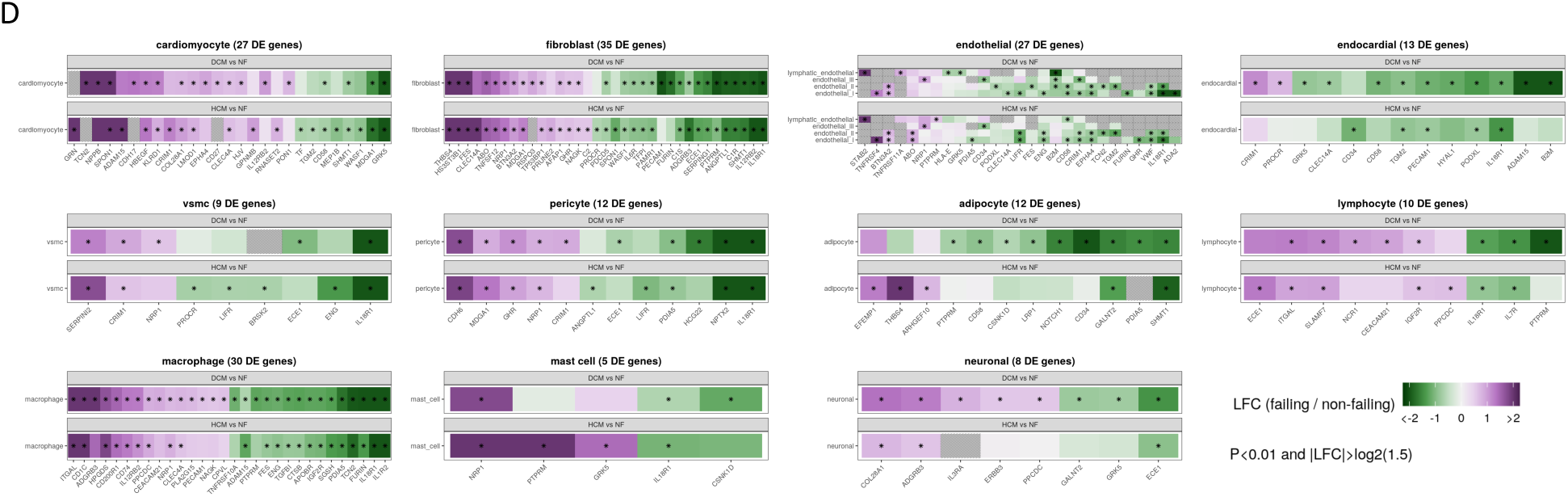
Enrichment of gene expression across different cell types in leP ventricle biopsies. A. Overall enrichment across 42 lel ventricle samples from 11 dilated cardiomyopathy (DCM), 15 hypertrophic cardiomyopathy (HCM) and 16 non-failing hearts combined. Proteins with significantly enriched gene expression in at least one cell type are shown. Average expression is scaled to center at 0 and capped to a range of -2.5 to 2.5 with a color gradient from yellow to purple. The percentage of nuclei expressing each gene is proportional to the size of the dots. The significant enrichment of gene expression (FDR_BH_-adjusted P-value < 0.01) in cell types are highlighted with black circles around the dots. Dots are omiQed for genes with less than 0.5% of nuclei within a cell type. B. Box plots of single-cell gene expression in log_2_ counts per million (CPM) of ADAM23, PAM and LPL in each cell type. FDR_BH_-adjusted *P*-values are reported only for genes in significantly enriched cell types. C. Box plots of single-cell gene expression (log_2_CPM) of NPPB and FURIN in non-failing, DCM and HCM heart samples stratified by cell types. Red asterisk highlights genes that are significantly differentially expressed in failing vs non-failing heart 3ssues. D. Differential gene expression in DCM or HCM *vs* non-failing heart samples stratified by cell types. The log_2_ fold change (LFC) of gene expression is scaled to center at 0 and capped to a range of –2 to 2, corresponding to a range of 0.25 to 4 3mes difference in gene expression. The significant difference of gene expression is labeled with asterisk in the square box.

Next, we investigated whether our prioritized targets were differentially expressed in cardiomyopathy and healthy donor heart biopsies in a cell-type specific manner. Indeed, 102 proteins (43.4%) were differentially expressed at the transcriptional level in at least one cardiac cell type (Figure 5D). *NPPB* showed significantly higher expression in cardiomyocytes from DCM, but not HCM, than non-failing hearts (Figure 5C). FURIN, mediating proteolytic cleavage of non-functional proBNP to its active hormone BNP, expressed at a significantly lower level in fibroblasts, endothelial cells and macrophages from DCM than non-failing hearts, suggesting agonists of FURIN may help restore cardioprotective activity of BNP in DCM.

## Discussion

### Main Findings

Leveraging genetic IVs to infer causalities between circulating proteins and incident diseases in a proteome-wide and phenome-wide manner help to identify candidate drivers of disease and biomarkers at an unprecedently large scale. In this study, we integrated proteomics MR with single cell analysis to prioritize novel targets for CVD. We found 221 causal signals, the majority of which conform to known disease biology through pathways such as immune response and atherosclerotic lesion formation, angiogenesis and vascular remodeling, myogenesis and cardiac progenitor cell differentiation, and energy metabolism. Reverse-MR identified 16 biomarkers whose protein expressions were affected by disease status, 5 of which were exclusively found in the East Asian population, demonstrating the value of multi-ancestry populations in identifying drug targets via genetic studies of molecular traits (omics). Only two of the targets were also biomarkers, the LGALS4 and the MMP12, suggesting distinct causes and consequences of circulating proteins on CVD. Some established targets for cardiovascular drugs approved or in clinical development have been found, such as PCSK9 inhibitors (bococizumab, alirocumab and evolocumab), ECE1 inhibitor (daglutril) and ANGPTL3 inhibitor (evinacumab), supporting the validity of our approach. About half of the candidate causal genes (112/235) have been linked to CVD or CVD-related risk factors in previous GWAS, genetic linkage or clinical studies, the rest of genes are implicated with CVD for the first time. Among the novel candidate causal genes to highlight, AGER, a member of the immunoglobulin superfamily of cell surface molecules, was linked to pathogenesis of atherothrombotic diseases, of which upregulated expression was found in human atherosclerotic plaques and polymorphisms associated with myocardial infarction and ischemic stroke;^45^ SELE, the E-selectin, and SELPLG, a glycoprotein ligand that binds to the E-, P- and L-selectin, mediating recruitment of leukocytes on vascular surfaces during initial steps of inflammation, were shown to be involved in development of atherosclerosis and thrombosis with increased plasma protein levels observed in various CVD;^46^ MDK, a secreted heparin-binding growth factor that regulates many biological processes, including cell proliferation, differentiation, migration and survival, was shown to have cardioprotective effects against ischemia and reperfusion injury via auenuating cardiomyocyte apoptosis^.47^ Our findings provide evidence supporting therapeutic hypotheses underpinning 3 established or under investigated cardiovascular drug targets and more broadly showcase the potential of large-scale integration of multi-omics in understanding causal human biology of complex disease for novel drug target discovery.

### The values and limitations of bidirectionality in MR analyses

Observational studies found associations without a capability to distinguish causal and confounding and are often lack of ascertainment of directionality of associations. We applied a bi-directional MR approach to disentangle causal roles of circulating protein level changes on disease risk – to discover targets, and changes on proteomic expression profiles under disease circumstances – to identify biomarkers. Distinct proteomic signatures were described as causes or consequences of different CVD, however, we were unable to distinguish disease incidence and progression, and thereby to assess proteomic changes underpinning disease progression due to limitations on disease outcome definitions from genome-wide meta-analyses. To achieve the full potential of bi-directional MR and characterize dynamic influence of proteomic changes along disease trajectories, a longitudinal GWAS on progression endpoints will be needed.

MMP12 is a notable example, of which the mechanism of action can be further elucidated with the longitudinal data. It was reported that MMP12 was upregulated after myocardial infarction, and then auenuated by endogenous inhibitors, such as tissue inhibitor of metalloproteinases (TIMPs) that provides a negative feedback loop to regulate a temporal succession of events that promote myocardial wound healing while limiting tissue damage^.48^ MMP12 levels are temporally fine-tuned after myocardial infarction, orchestrating a series of events including inflammation, fibrosis, angiogenesis and collagen degradation in order to achieve an optimal scar formation and prevent left ventricular dysfunction and heart failure prognosis.^49^ In our findings, we confirmed that MMP12 was a biomarker that increased expression after myocardial infarction, and activation of MMP12 reduced risk of peripheral arterial disease and ischemic stroke, but because of limitations on data availability of the exact time point when MMP12 protein level was quantified after ischemic injury, we were not able to assess the temporal regulation of MMP12 during cardiac remodeling upon the injury. Further studies that include disease progression endpoints can help corroborate and extend our findings to characterize dynamic protein level changes of cardiovascular targets and biomarkers.

### Concordance and differences of using *cis-* and *trans-*pQTLs as IVs in MR

Using *cis-* and *trans-*pQTLs as IVs represent fundamentally distinct principles of IV selection, thereby affecting interpretation of the forward-MR results. *cis-*pQTLs are physically proximal to sentinel variants, and therefore less likely to violate the MR assumption of pleiotropy, while *trans*-pQTLs include more IVs that provide greater statistical power and the IVs representing genes that may have biological roles in regulating homeostasis of the associated proteins through signaling transduction pathways or protein-protein interaction.^50^ Forward-MR identified 221 proteins that showed significant causal effects on at least one CVD, among which 59 are common targets identified using either *cis-* or *trans-*pQTLs. One hundred twenty-nine (68.6%) targets were uniquely identified in the *trans-*pQTLs-based MR analysis, accounting for 81.7% of targets with biological annotations that implied a pathophysiological role in CVD, suggesting the value of using *trans*-pQTLs in the MR analysis for exploring novel therapeutic hypotheses. Functional follow-up analyses are needed for validating *trans*-pQTLs derived targets. On the contrary, some targets are found only in the *cis-*pQTLs-based MR analysis, for example, NOTCH1 for cardiac valvular disease. NOTCH1 is a Notch receptor that releases its intracellular domain as transcription factor upon activation, which plays an essential role in cell fate determination, and cell proliferation, differentiation and apoptosis during organogenesis throughout the embryo.^51^ Loss-of-function mutations in NOTCH1 was found to cause aortic valve disease in autosomal-dominant human pedigrees that showed a wide spectrum of developmental aortic valve anomalies and severe valve calcification.^52^ Agonists of NOTCH1 may be able to treat calcification of the aortic valve, the third leading cause of CVD in adults. Taken together, proteomic MR with *cis-* and *trans-*pQTLs provides complementary approaches for therapeutic target identification with overlapped but unparalleled insights.

### Impact of applying different methods for multiple testing correction

The forward-MR generated more targets that reached the Bonferroni-corrected significance threshold than biomarkers from the reverse-MR. We noted that Bonferroni-correction that corrected for the total number of proteins and phenotypes analyzed in the phenome-wide MR scan can be overly stringent due to intercorrelation within proteins and phenotypes. Using the Bonferroni approach controlled type 1 error with a slight loss in power, given the relatively low success rates for investigational targets passing through clinical trial development,^53^ the strict control on false positives at the first step of target discovery can help prioritize targets for downstream development. If using a more lenient approach to adjust for multiple testing, such as the Benjamini–Hochberg method of false discovery rate, we found the number of significant proteins for forward-MR increased from 221 (7.52%) to 1,349 (69.54%), for reverse-MR from 16 (0.54%) to 1,045 (35.54%), while resulting in a larger number of significant findings, more of which can be false positives that require extensive functional appraisal with labor-intensive and time-consuming experiments developed on animal models. External validation using independent cohorts can validate our results on top of a stringent significance threshold applied, however, at the time when this manuscript is wriuen up, the pQTLs we used are from the largest proteomic GWAS recently performed under the UK Biobank Pharma Proteomics Project.^19^ Future validation is required when pQTLs from equally-powered, independent studies are available.

### Benefits and limitations in incorporating multi-ancestry element in MR study

Leveraging multi-ancestry data for drug discovery can help discover novel candidate causal genes. For example, two nonsense mutations of PCSK9 first reported in humans were found in an African-American population and associated with a substantial reduction of low-density lipoprotein cholesterol.^54^ This finding demonstrated that a lifelong inhibition of PCSK9 protected against coronary heart disease without noticeable safety issues. Multi-ancestry element has recently been increasingly incorporated in population genetic studies, such as GWAS, fine-mapping and polygenic risk score studies, but rarely applied in MR studies.^55,56^ In our study, we identified 5 biomarkers exclusively detectable in the East Asian population, highlighting the values of leveraging multi-ancestorial populations in identifying candidate causal signals. We benefited from the relatively large sample size of the Biobank Japan, one of the largest non-European population cohorts with genome-wide genetic and medical records data available, which provided adequate statistical power in constructing IVs of disease endpoints and thereby detecting significant signals in the reverse MR, while hindered from the relatively small sample size of the pQTL study in this population, limiting us from identifying more potential target signals in the forward MR.

### Conclusions

Our study evaluated causal relationships between 3,000 circulating proteins and 21 CVD using biobank-scale genetic association summary statistics for proteins and phenotypes in a bi-directional MR framework. We prioritized novel candidate causal signals for CVD and interrogated their pleiotropy and specificity in a phenome-wide causality survey. This study provides human genetics-based evidence of novel candidate genes, a foundational step towards full-scale causal human biology-based drug discovery for CVD.

## Data Availability

All data produced in the present study are available upon reasonable request to the authors.

